# Brain Death versus Circulatory Death: How Functional Warm Ischemia and Cold Storage Shape Myocardial Repair and Damage in Human Donor Hearts

**DOI:** 10.1101/2025.09.08.25335328

**Authors:** Shiyi Li, Rishav Bhattacharya, Abdussalam E. Elsenousi, Katherine V. Nordick, Adel M. Hassan, Syed B. Peer, Camila Hochman-Mendez, Todd K. Rosengart, Kenneth K. Liao, Nandan K. Mondal

## Abstract

This study compares myocardial injury responses in human donor hearts from donation after brain death (DBD) and donation after circulatory death (DCD), with a focus on myocardial membrane integrity, pyroptosis, and damage. Unlike DCD hearts, which are exposed to varying durations of functional warm ischemic times (fWITs), DBD hearts - never subjected to warm ischemia - served as controls. A total of twenty-four human hearts were procured, consisting of six from the DBD group and eighteen from the DCD group. All procured hearts were placed in cold normal saline and stored for up to six hours. Left ventricular biopsies were performed at 0, 2, 4, and 6 hours to assess plasma membrane repair proteins (Annexin A1, Dysferlin), pyroptosis markers (NLRP3, caspase-1, GSDMD-NT), and to evaluate edema and injury scores. Data suggest that DBD hearts maintained stable levels of plasma membrane repair proteins and showed no evidence of pyroptosis activation or significant injury throughout cold storage. In contrast, DCD hearts exhibited profound Annexin A1 depletion, early and progressive pyroptosis, elevated edema, and worsening histopathological injury - directly correlated with fWITs. These findings underscore that warm ischemia is a critical determinant of pyroptotic damage in donor hearts, and highlight the relative resistance of DBD hearts to such injury during preservation. For DCD hearts, strategies to enhance membrane repair capacity and inhibit pyroptosis should focus on the fWIT phase to assess donor heart quality and suitability for transplantation.

**New & Noteworthy:** This study demonstrates that donor hearts procured after circulatory death (DCD) exhibit early Annexin A1 depletion and activation of the NLRP3/caspase-1/GSDMD-mediated pyroptosis pathway during cold storage – a phenomenon absent in brain-dead (DBD) donors. We establish a direct correlation between warm ischemia time and pyroptotic damage in DCD hearts. These findings identify Annexin A1 as a key mediator of ischemia injury and a promising therapeutic target to improve viability in marginal donor hearts.

## Introduction

Despite significant advances in mechanical circulatory support, heart transplantation using brain death donor (DBD) hearts remains the gold standard therapy for end-stage heart failure patients, yet it is limited due to the shortage of DBD donors.(1) Recent clinical experiences across multiple countries have demonstrated that the adoption of hearts from donation after circulatory death (DCD) can substantially increase the donor pool and reduce waitlist mortality.(1, 2) Moreover, a large multi-center randomized trial reported that risk-adjusted survival at six months following DCD heart transplantation was non-inferior to that of donation after brain death (DBD) heart transplantation.(3) Over the past decade, DCD heart transplantation has grown rapidly.(4) However, its broader clinical application continues to face three major challenges: the inevitable effects of warm ischemia injury, the lack of a cost-effective preservation strategy, and unresolved ethical concerns.

For DCD liver and kidney, static cold storage remains the primary preservation method for transplantation in many centers.(5, 6) Cold static storage offers a simple and cost-effective method for DBD heart preservation. However, its use in DCD heart preservation has been limited due to concerns of rapid deterioration in function during cold storage. Understanding the impact of cold ischemia injury on DCD hearts may offer valuable insights into pharmacological interventions that could optimize cold preservation solutions for DCD hearts. This preservation strategy may present a viable option for clinicians in specific circumstances, such as short-distance donor heart transportation and resource-limited settings.

In our previous study, we demonstrated that prolonged cold storage increases the susceptibility of DBD hearts to ferroptotic cell death. In contrast, warm ischemic injury has been identified as the primary factor that causes the vulnerability of DCD hearts to ferroptosis, a distinct mechanism of regulated cell death.(7) Plasma membrane disruption is not merely a symptom but a central executing event in ischemic irreversible cell death.(8) The failure of energy-dependent pumps causes physical and enzymatic damage that ruptures the membrane, which in turn directly causes the lethal loss of homeostasis, triggers destructive inflammatory pathways, and activates another regulated cell death mechanism, the pyroptotic cell death. The current study aimed to identify this process as another core mechanism differentiating the injury in DCD hearts (which experience warm ischemia) from DBD hearts (which do not) under cold storage conditions.

## Materials and Methods

### Donor enrollment for research

For this study, we obtained human hearts initially offered for DBD or DCD heart transplantation but were subsequently turned down for various reasons, such as age, size and weight mismatch, distance, and logistics. Our institutional IRB has determined that our research, which used the hearts from circulatory death and brain death donors rejected for transplantation, does not qualify as a human study and thus does not require IRB approval. Our institution has a research agreement with the local Organ Procurement Organization (Life Gift, Houston, TX), which allows us to obtain donated human hearts that are authorized and approved for medical research.

### Inclusion and exclusion criteria for research

Hearts were procured from donors aged 18 years or older. To be included, donors must have had no documented history of coronary artery disease, prior cardiac surgery, active cancer, significant troponin elevation, or known cardiac contusion.

Hearts were excluded from the study for the following reasons:

- A history of coronary artery disease or previous cardiac surgery.
- An abnormal electrocardiogram (ECG).
- Evidence of abnormal cardiac function on echocardiogram.
- For donors after circulatory death (DCD), failure to expire within 90 minutes following withdrawal of life-sustaining therapy.

Additionally, no protected health information was accessed prior to the declaration of donor death.

### Donor heart procurement and study design

This study cohort consisted of 24 human donor hearts with a median age of 45 years (range: 18– 66) and included 58% male donors. Among these, 6 were obtained after brain death (DBD) and 18 after circulatory death (DCD). The DBD hearts served as the control group, as they were not exposed to warm ischemia. Following cross-clamp, one liter of del Nido cardioplegia (Nephron Pharmaceuticals Co., West Columbia, SC, USA) was administered. The procured hearts were stored in a single container with 1 L of cold normal saline and maintained on ice in a cooler for up to 6 hours. For DCD donors, functional warm ischemia time (WIT) was defined as the interval from systolic blood pressure dropping below 50 mmHg until cardioplegia induction with del Nido. Left ventricular (LV) biopsies were collected from all hearts at baseline (0 hours) and after 2, 4, and 6 hours of cold static storage. All tissue samples were immediately snap-frozen in liquid nitrogen for subsequent analysis. A schematic overview of the procurement, storage, and experimental timeline is provided in **Figure 1**.

**Figure 1.**
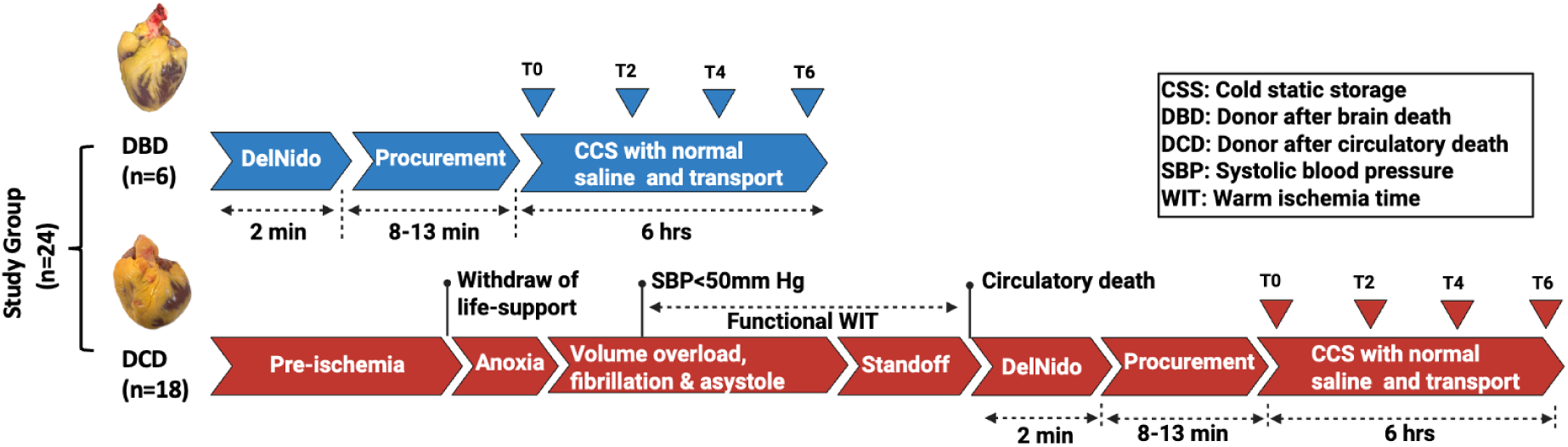
Schematic representation of the process and timelines for the DBD and DCD donor heart procurements and storage for research. Note: DBD, donor after brain death; DCD, donor after circulatory death; WIT, warm ischemia time; CSS, cold static storage; SBP, systolic blood pressure; T0, cold static storage time at baseline; T2, T4, T6, cold static storage time at hours 2, 4, and 6, respectively.

### Preparation of myocardial protein

Frozen LV tissue samples were homogenized in ice-cold RIPA buffer (Thermo Fisher Scientific, Cat. No. 89901) containing PMSF (Cell Signaling, Cat. No. 8553) and protease inhibitor cocktail (Thermo Fisher Scientific, Cat. No. 78439). The extract was centrifuged at 12,000 g for 15 minutes at 4°C, and the supernatant was aliquoted and kept in the freezer (−86°C) until further use. Myocardial protein concentrations were measured by Quick Start Bradford Protein Assay (Bio-Rad, Cat. No. 5000201), following the manufacturer’s instructions.

### SDS-PAGE and Western blotting

Sample preparation was carried out in Laemmli buffer using an equal amount of cardiac protein, following our previously published protocol.(7, 9) The protein was then run on a 4–20% Mini-PROTEAN® TGX™ Precast Protein Gel (Bio-Rad, Cat No. 4561094) at a constant current until the dye reached the bottom. After electrophoresis, a methanol-activated polyvinylidene difluoride (PVDF) membrane (Bio-Rad, Cat No. 1620174) was used for protein transfer. To prevent non-specific binding of antibodies, the membrane was blocked with 5 % (w/v) bovine serum albumin (BSA: Cell Signaling, Cat No. 9998s) prepared in tris-buffered saline (TBS) containing 0.1 % (v/v) Tween 20 (TBS-T) for 60 minutes at room temperature. Primary antibody incubation was conducted overnight at 4°C. Following this, the membrane was washed with TBS-T six times at 5-minute intervals to remove any unbound primary antibody. After washing, the membranes were exposed to a secondary antibody [horse radish peroxidase-conjugated goat anti-rabbit IgG (H+L), 1:5000 dilution, Invitrogen, Cat No. 31460, RRID: AB_228341] for 60 minutes at room temperature. Subsequently, the membranes were washed and developed with Pierce ECL Western Blotting Substrate (Thermo Scientific, Cat No. 32109). The membranes were visualized using the Odyssey® Fc Imaging System (LI-COR Biosciences, Nebraska, USA, RRID: SCR_023227). To detect membrane repair proteins, we utilized the following primary antibodies: Annexin A1 (dilution 1:1000, Cell Signaling, Cat No. 3299, RRID: AB_2274094) and Dysferlin (dilution 1:1000, Abcam, Cat No. ab124684, RRID: AB_10976241). The total protein expression was measured and then normalized using the loading control GAPDH (dilution 1:2000, ThermoFisher Scientific, Cat No MA5-15738, RRID: AB_10977387). Afterward, the same membranes were stripped using ReBlot Antibody Stripping Solution (Millipore Sigma; Cat No. 2504) and probed for the corresponding loading control. The density of each band was normalized with the respective GAPDH density using UN-SCAN-IT gel software (Silk Scientific, Inc., Utah, USA, RRID: SCR_017291).

### Enzyme-linked Immunosorbent Assay

We used ELISA kits to measure the levels of human cardiac Annexin A1 (My BioSource; Cat No. MBS761416), Dysferlin (My BioSource; Cat No. MBS2024255), and N-terminal proteolytic fragment of Gasdermin D (GSDMD-NT) (My BioSource; Cat No. MBS7273502) following the manufacturer’s instructions.

### Tdt-mediated dUTP nick end labeling (TUNEL) Assay

TUNEL assay to detect DNA fragmentation of individual cells was conducted using the in-situ cell death detection kit (Roche; Cat No. 11684795910) following the manufacturer’s instructions. The nuclei were stained with DAPI (1 µg/ml) for 5 min at room temperature. TUNEL-stained slides were imaged under a EVOS M7000 microscope (EVOS M7000 Imaging System, ThermoFisher Scientific, Waltham, MA, RRID: SCR_025070) in 6 fields of view. Nuclei that were double-labeled with DAPI and TUNEL were considered positive. The number of positive cells in relation to the total number of cells was quantified in tissue sections.

### Assessment of Myocardial Edema

To detect myocardial edema, the water content of heart tissues was measured by the wet and dry weight method according to our previously published procedure(10). Briefly, at baseline, at hours 2, 4, and 6, a small portion of left ventricular tissue samples from the donor’s heart was weighed to obtain wet weight (WW). The tissue samples were then dried in an oven at 80°C for 72 h and weighed again to get the dry weight (DW) at four different time points for each heart. The formula (WW−DW)/WW × 100% was used to calculate myocardial edema and expressed as a percentage of wet weight.

### Routine Histology: Hematoxylin & Eosin Staining

Myocardial tissue slices were fixed in 4% paraformaldehyde solution (Thermo Fisher Scientific, Cat No. J19943.K2), embedded in paraffin, cut to 4-μm-thick sections, and stained with commercially available Hematoxylin and Eosin (H&E) kit (Abcam, Cat No. ab245880). Histological scoring of the H&E-stained slides was performed, followed by our previously reported procedure.(10)

### Immunohistochemical Analysis of Caspase-1 Expression

To evaluate myocardial caspase-1 expression, immunohistochemical (IHC) staining was performed using a mouse and rabbit-specific HRP/DAB (ABC) detection kit (Abcam, Cat. No. ab64264) in accordance with the manufacturer’s protocol. Tissue sections were incubated overnight at 4 °C with a primary antibody directed against caspase-1 (Abcam, Cat. No. ab62698, RRID: AB_955727) at a dilution of 1:100. Immunostaining results were quantified using a previously established scoring methodology.(11)

### Statistical analysis

All statistical analyses were performed using GraphPad Prism (version 9.5.1, GraphPad Software, Inc., La Jolla, CA, RRID: SCR_002798) and SAS software (version 9.4, SAS Institute, Cary, NC, RRID: SCR_008567). Continuous variables, expressed as median with interquartile range (IQR), were compared between donor groups using the non-parametric Kruskal–Wallis test (one-way ANOVA), while categorical variables, presented as counts and percentages (n, %), were compared using Fisher’s exact test. Heatmap and principal component analysis (PCA) plots were generated using matplotlib version 3.10.0 (RRID: SCR_008624) and scikit-learn version 1.6.1 (RRID: SCR_002577) in Python version 3.12.9 (RRID: SCR_008394). Ellipses were drawn on the scatterplots for 90% confidence intervals. A p-value of less than 0.05 was considered statistically significant.

## RESULTS

### Demography of donor: DBD versus DCD

The demographic and clinical characteristics of the DBD and DCD groups are summarized in **Table 1**. The cohorts were broadly comparable across all measured parameters. The median donor age was similar between groups (DBD: 41 years [IQR 26–53] vs. DCD: 44 years [IQR 40–51]; p = 0.640). There was a higher proportion of male donors in the DCD group (67.67%) compared to the DBD group (33.33%), though this difference was not statistically significant (p = 0.192). Racial distribution differed somewhat, with the DBD group consisting predominantly of Hispanic donors (50%), whereas the DCD group had a higher proportion of White donors (61.11%). The majority of donors in both groups were obese, with median BMI values exceeding 30 kg/m². Substance abuse history, including smoking and alcohol use, was more common among DCD donors, but no significant differences were found in substance use, ABO blood group distribution, cause of death, or echocardiographic parameters such as left ventricular ejection fraction between the two groups. These results confirm that the DBD and DCD donor cohorts were well-matched at baseline.

**Table. 1:**
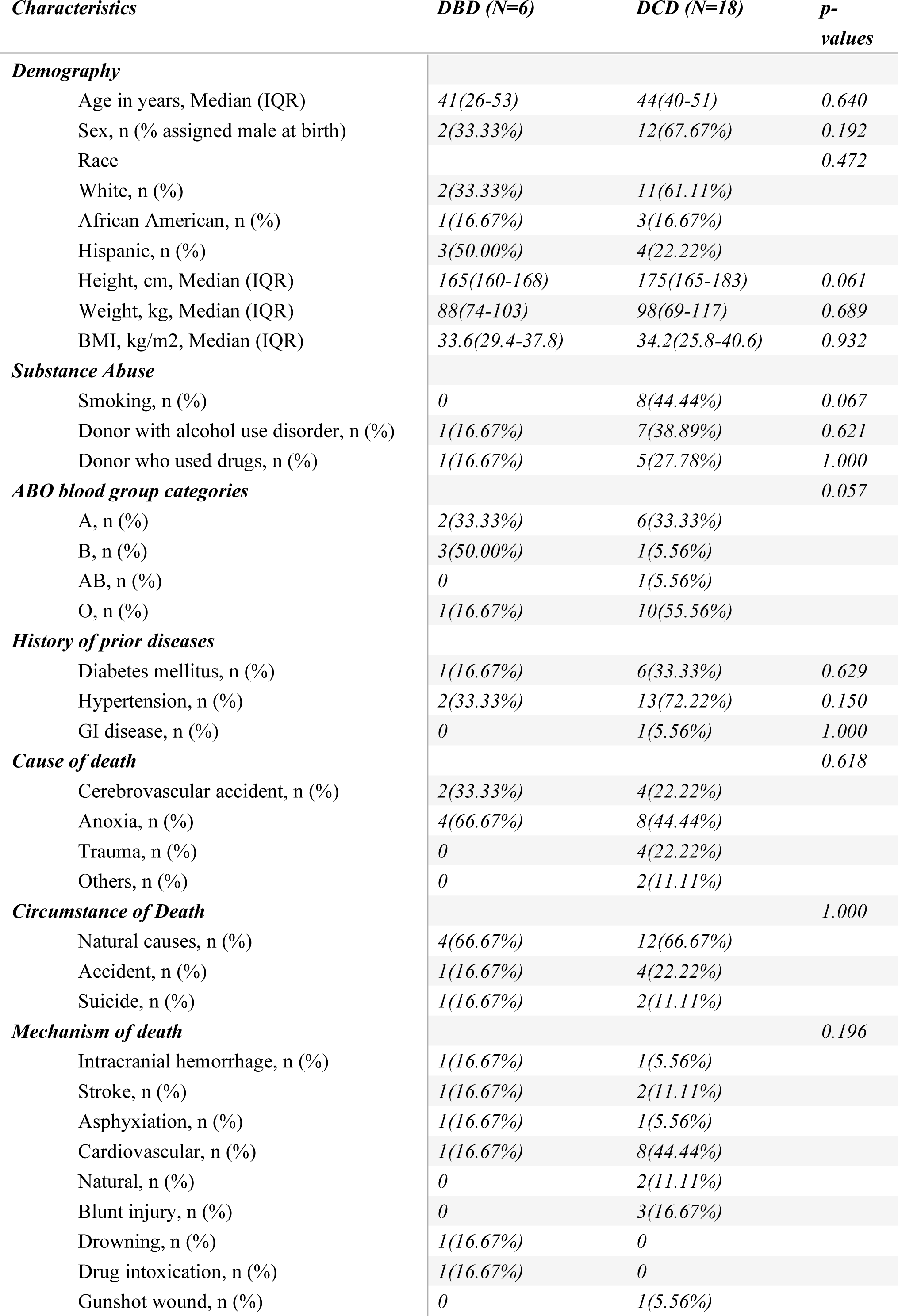
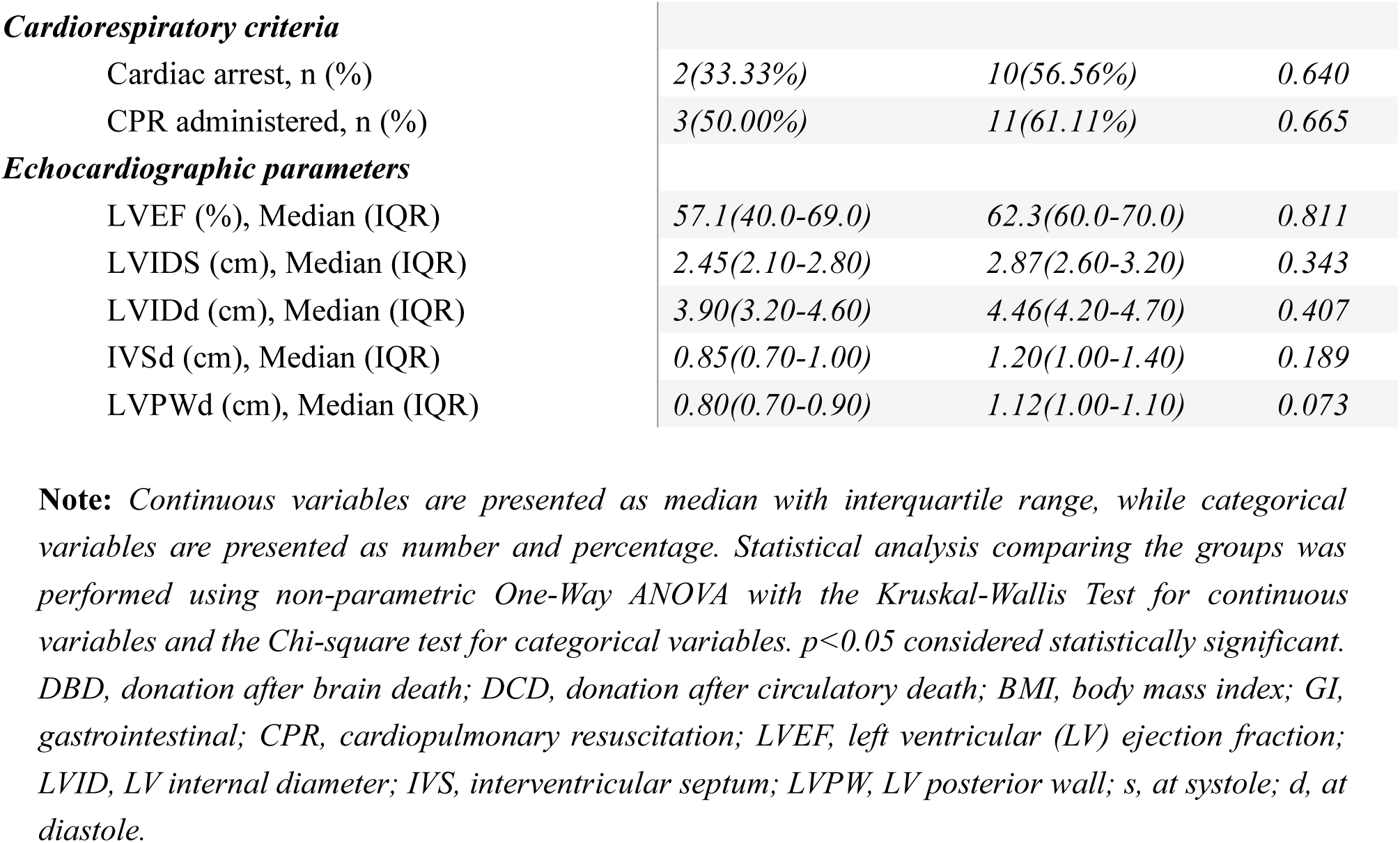
Demographic and baseline clinical characteristics of DBD and DCD donors.

### Annexin A1 Deficiency Drives Membrane Disruption in DCD but Not DBD Hearts

A stark contrast in plasma membrane integrity was observed between DBD and DCD hearts, driven primarily by the depletion of Annexin A1 (**Fig.2**). At baseline, prior to cold storage, DCD hearts already exhibited significantly lower levels of Annexin A1 compared to DBD hearts, as confirmed by both immunoblot and ELISA analyses (**Fig.2A,B,C**). In contrast, Dysferlin expression remained comparable between the two groups at this initial time point (**Fig.2A,D,E**). Throughout the six-hour cold storage period, Annexin A1 levels showed a slight increasing trend in DBD hearts but declined progressively in DCD hearts, resulting in consistently lower Annexin A1 in DCD samples at all time points (**Fig.2F,G**). Dysferlin expression, however, remained stable over time and did not differ significantly between DBD and DCD groups (**Fig. 2A,H,I**).

**Figure 2.**
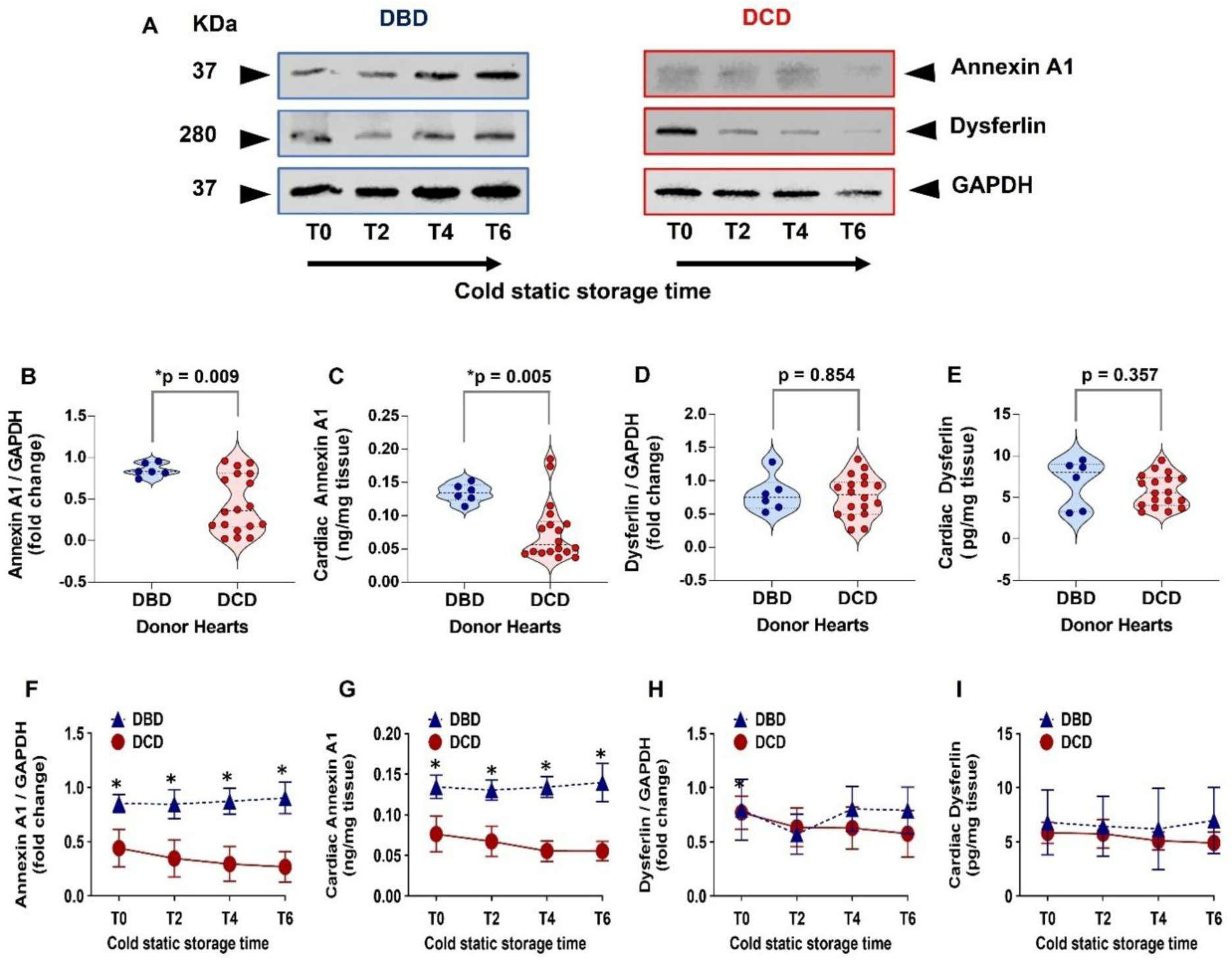
(A) Representative immunoblots of Annexin A1, Dysferlin, and corresponding loading control (GAPDH) between DBD and DCD hearts under multiple time points of cold static storage; (B-E) Violin plots for semi-quantitative immunoblot and ELISA quantification for (B,C) Annexin A1 and (D,E) Dysferlin in DBD and DCD groups at baseline. (F-I) Temporal changes in semi-quantitative immunoblot and ELISA quantification between DBD and DCD for **(F,G)** Annexin A1 and **(H,I)** Dysferlin protein expression assessed over 6 hours of cold storage. Note: *p < 0.05 is considered significant. DBD, donor after brain death; DCD, donor after circulatory death.

### Pyroptosis Signaling Escalates During Cold Storage in DCD But Not DBD Hearts

Quantitative ELISA data for cardiac pyroptosis markers NLRP3 and immunohistochemically detected caspase-1 in DBD and DCD hearts are presented in **Figure 3**. DCD hearts exhibited significantly higher levels of NLRP3 and caspase-1 at baseline compared to DBD hearts (**Fig.3A,C**). Throughout cold storage, this divergence widened dramatically: NLRP3 and caspase-1 levels showed a sharp, progressive increase in DCD hearts, peaking at 6 hours, while levels in DBD hearts remained low and stable (**Fig.3B,D**). A panel of IHC images showing caspase-1 positive cells in DBD and DCD hearts at multiple timepoints (T0-T6) is presented in **Figure 3E(a–h)**. This indicates that warm ischemia primes DCD hearts for a robust and escalating pyroptotic inflammatory response during subsequent cold storage, a phenomenon entirely absent in DBD hearts.

**Figure 3.**
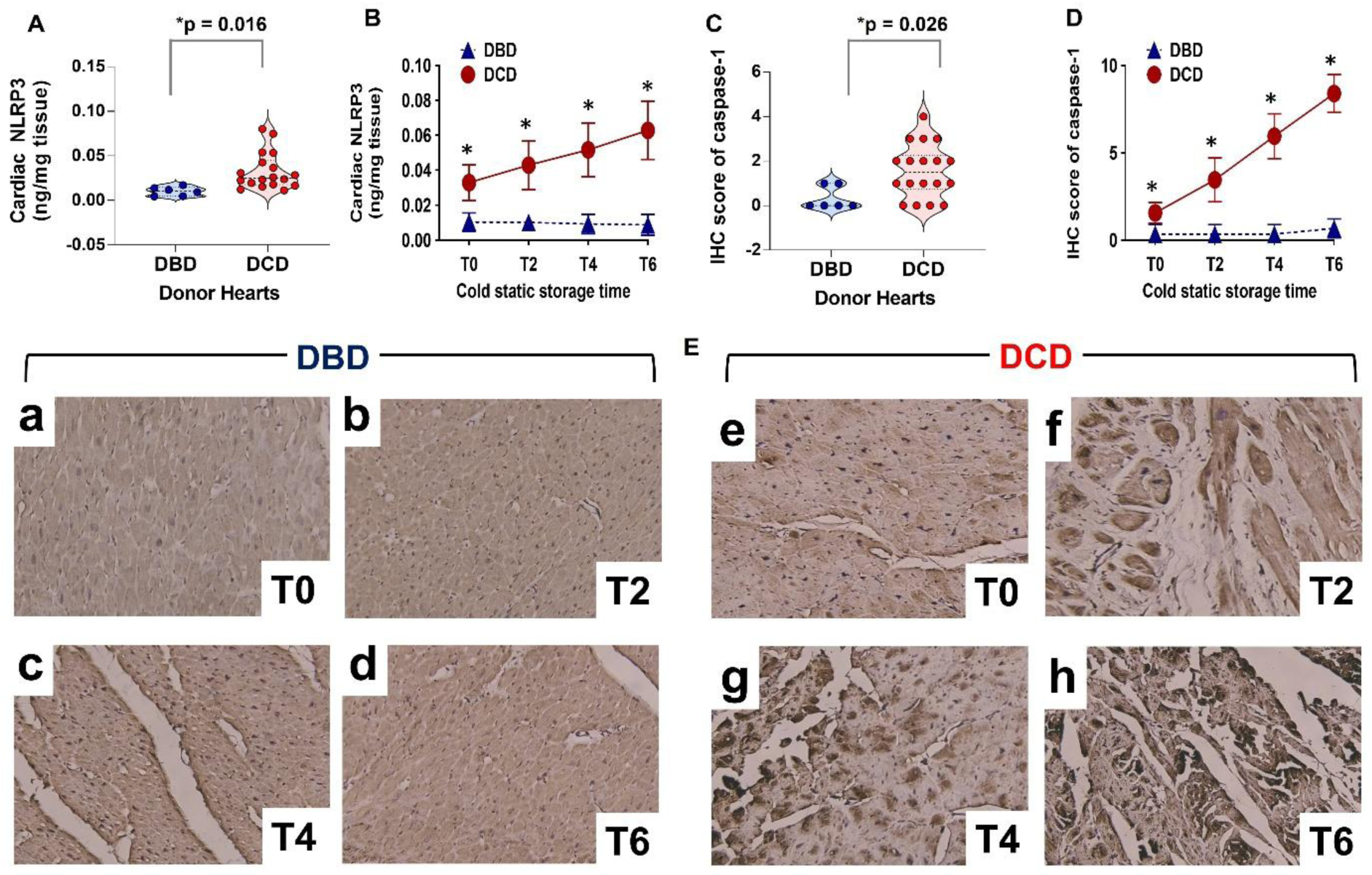
*(A-D) Violin plot and line arts for quantitative ELISA data for* pyroptosis markers *(A,B)* NLRP3, and immunohistochemical score of (C,D) caspase-1 at (A,C) baseline and (B,D) multiple timepoints after cold storage between DBD and DCD hearts. (E) Representative immunohistochemistry images of caspase-1 in DBD and DCD hearts at different time points of cold storage (a-h), magnification 160x. *Note: *p < 0.05 is considered significant. DBD, donor after brain death; DCD, donor after circulatory death*.

Quantitative analysis revealed that cleaved GSDMD-NT, the executioner protein of pyroptosis, was significantly elevated in DCD hearts at baseline and increased sharply over 6 hours of cold storage, whereas levels in DBD hearts remained consistently low and stable (**Fig.4A, B**). This inflammatory cell death was further reflected in TUNEL assays, where DCD hearts showed moderately higher baseline TUNEL-positive cells and a marked temporal increase, reaching over 50% after 6 hours, while DBD hearts maintained near-absent TUNEL positivity throughout preservation (**Fig.4C,D**). Representative immunofluorescence images visually corroborated the stark contrast in TUNEL staining between groups before and after cold storage (**Fig.4E(a–h)**). These results demonstrate that warm ischemia initiates irreversible pyroptotic cell death in DCD hearts, which is profoundly exacerbated by subsequent cold storage, a response not observed in DBD hearts.

**Figure 4.**
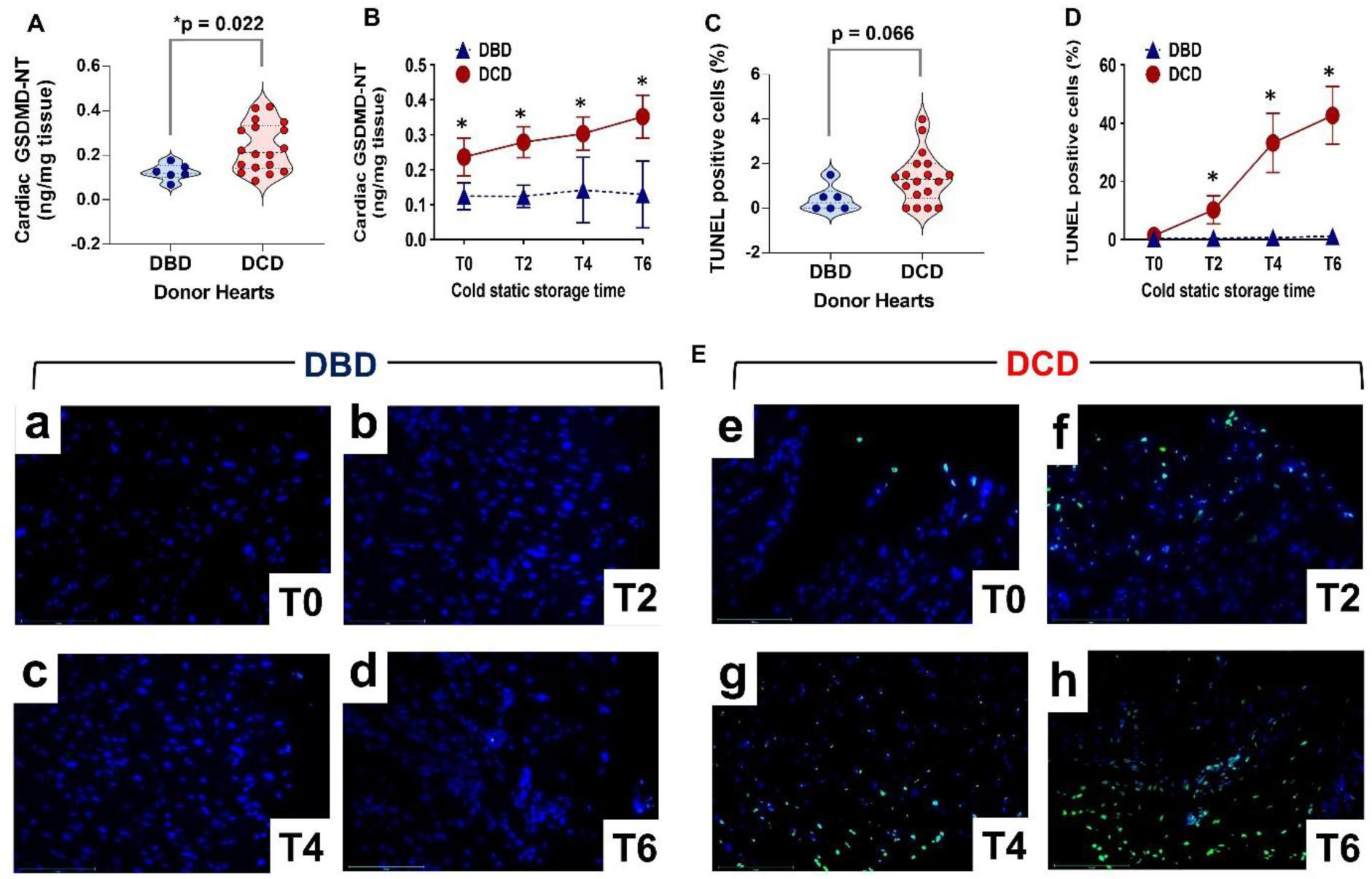
(A-D) Violin plot and line arts for quantitative ELISA data for pyroptosis markers (A,B) GSDMD-NT, and (C,D) TUNEL-positive nuclei at (A,C) baseline and (B,D) multiple timepoints after cold storage between DBD and DCD hearts. (E) Representative images showing merged staining (DAPI in blue, TUNEL in green) at various time points during cold static storage (a-h) between DBD and DCD hearts. The TUNEL-positive myocytes are expressed as a percentage of total cells counted. Note: *p < 0.05 is considered significant. DBD, donor after brain death; DCD, donor after circulatory death.

### Edema and Injury in DBD vs. DCD Hearts During Cold Storage

Myocardial water content (edema) and histopathological injury scores from DBD hearts and DCD hearts across multiple time points during cold static storage are presented in **Figure 5**. Prior to cold storage, DCD hearts exhibited significantly more severe edema (**Fig.5A**) and cardiac injury (**Fig.5B**) compared to DBD hearts. Following six hours of cold storage, both DBD and DCD groups showed a gradual temporal increase in myocardial edema and histopathology scores; however, the values in the DCD group consistently remained higher throughout the storage period (**Fig.5C,D**). Representative H&E-stained images of DBD and DCD hearts before and after cold storage are shown in **Figure 5E(a– h)**.

**Figure 5.**
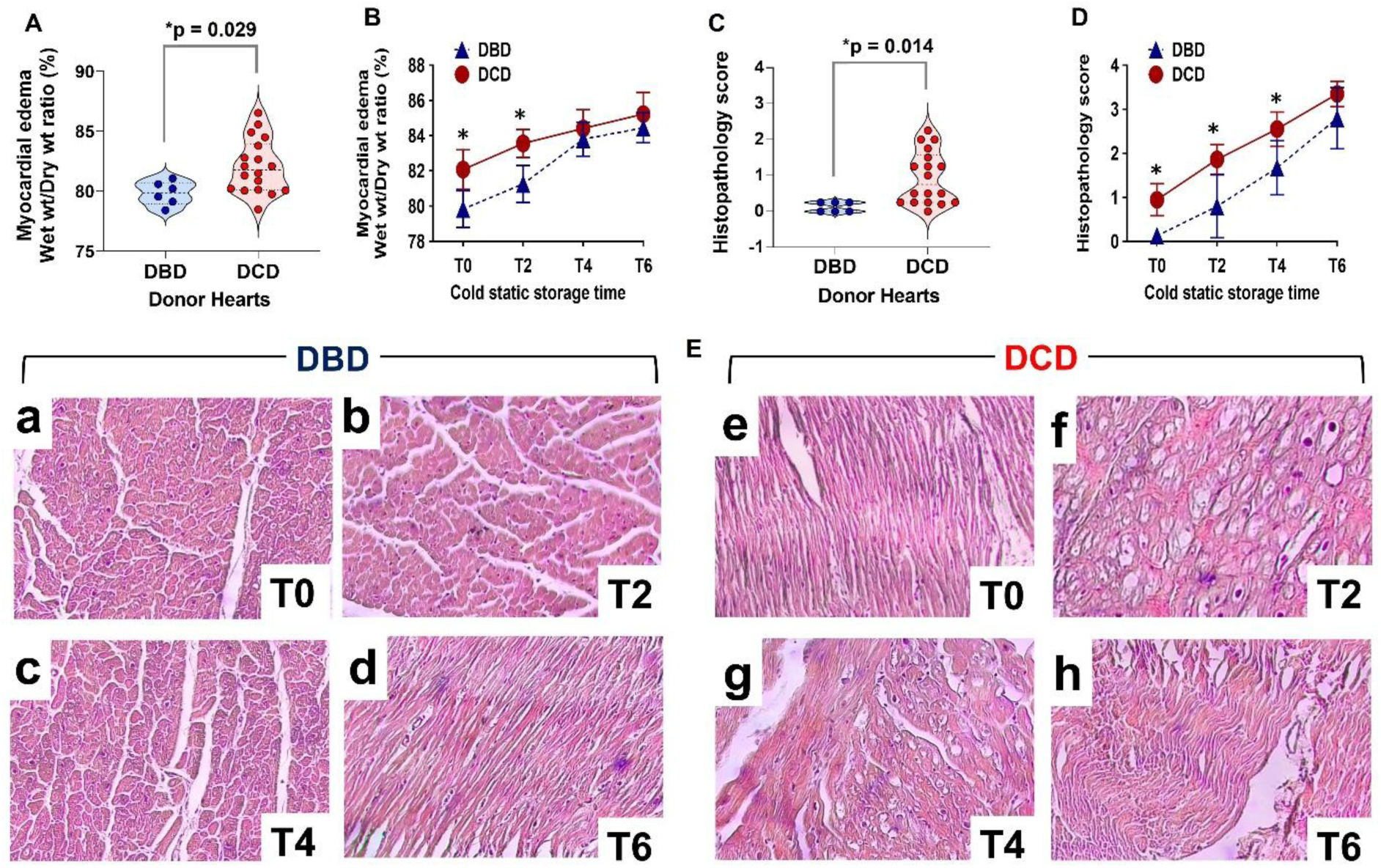
(A-D) Violin plot and line arts for (A,B) myocardial edema, and (C,D) histopathology injury score at (A,C) baseline and (B,D) multiple timepoints after cold storage between DBD and DCD hearts. (E) Representative H&E images showing injury score at various time points during cold static storage (a-h) between DBD and DCD hearts, magnification 160x. Note: *p < 0.05 is considered significant. DBD, donor after brain death; DCD, donor after circulatory death.

### Integrated Biomarker Profiling Reveals Warm Ischemia Time as the Primary Driver of Pyroptotic Activation and Cellular Damage in donor Hearts

A global analysis of biomarker expression, normalized by z-score and visualized via heatmap, revealed coordinated cellular changes across donor groups and preservation timepoints (**Fig.6A**). Annexin A1 was markedly deficient in DCD hearts, with the severity of depletion strongly correlated with longer Warm Ischemic Time (WIT). In contrast, Dysferlin expression showed no consistent pattern between DBD and DCD groups. Pyroptosis execution markers (NLRP3, GSDMD-NT) and cell death markers (caspase-1, TUNEL) were minimally active in DBD hearts but became significantly activated in DCD hearts, increasing with both prolonged cold storage and, more profoundly, with longer WIT. This pattern was mirrored in tissue-level injury, where edema and histopathology scores worsened with cold storage time in all hearts, but were severely exacerbated by WIT in DCD hearts.

**Figure 6.**
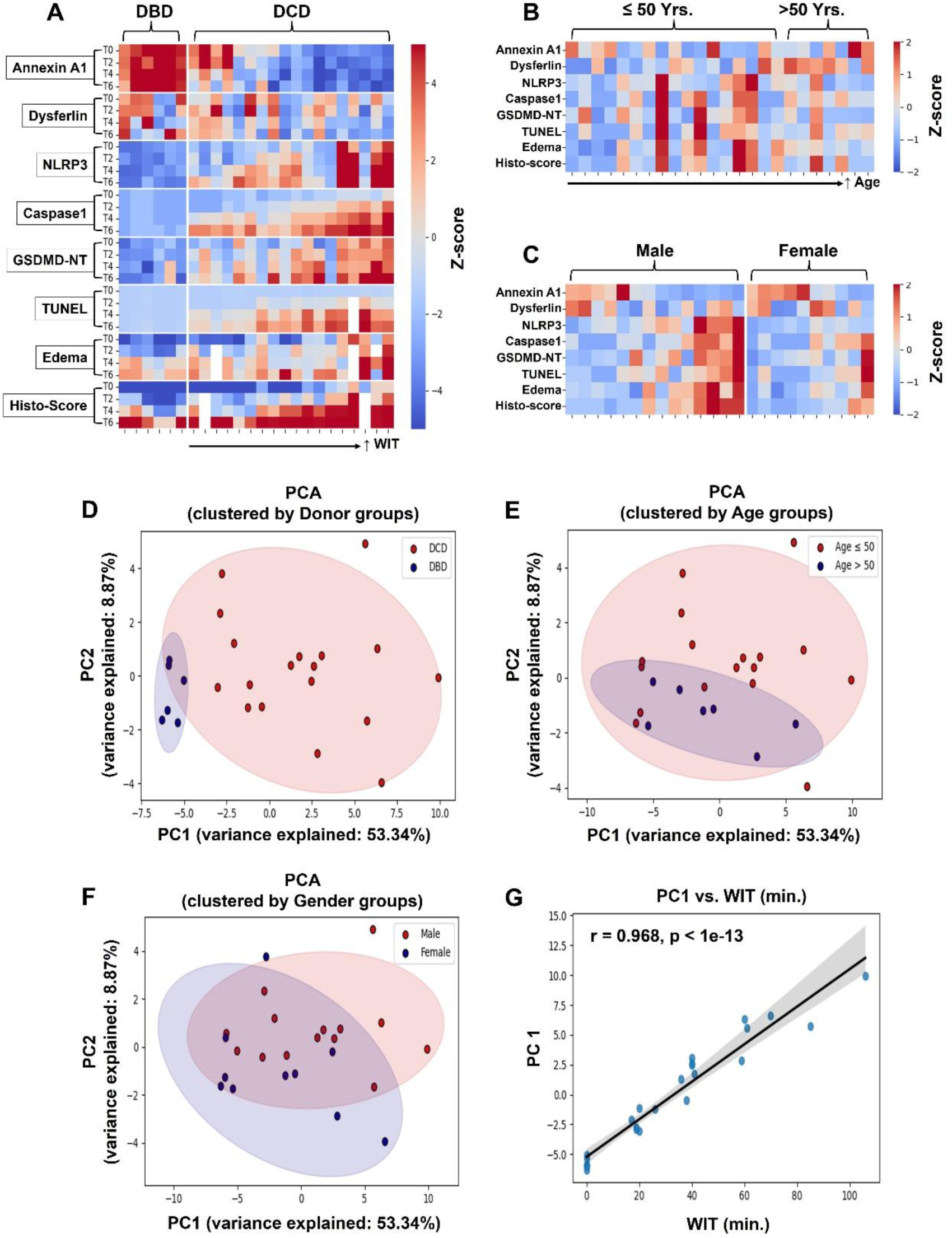
(A-C) Heatmaps and (D-F) PCA scatterplots showing all biomarker changes in concert. (A) the biomarker values for all patients at all time points, while (B,C) highlight the changes at baseline in relation to (B) Age and (C) gender. (D-F) the same PCA plot, clustered into different groups (90% confidence ellipsis), with (D) DBD vs DCD, (E) age ≤ 50 vs age > 50, and (F) male vs female. (G) a linear regression between WIT on the x-axis and PC1 embeddings on the y-axis. Note: DBD, donor after brain death; DCD, donor after circulatory death; PCA, principal component analysis; WIT, warm ischemic time in minutes.

The baseline biomarker profile was not significantly influenced by donor age (**Fig.6B**) or gender (**Fig.6C**). Principal Component Analysis (PCA) reduced the biomarker data into two components, with PC1 (53.34% of variance) and PC2 (8.7% of variance) (**Fig.6D**). The clusters for DBD and DCD hearts showed clear separation along PC1, while stratification by age and gender resulted in weaker separation primarily along PC2 (**Fig.6E,F**). The dominant separation between DBD and DCD groups was driven by PC1, which demonstrated a near-perfect correlation with Warm Ischemic Time (r = 0.968, p < 1e-13) (**Fig.6G**), unequivocally identifying WIT as the foremost factor governing the divergent molecular and injury responses observed.

## 4 Discussion

The critical shortage of donor hearts has necessitated the exploration of DCD heart transplantation as a viable strategy to expand the donor pool. While outcomes are promising, the inevitable period of fWIT remains a significant barrier to its widespread adoption, particularly when considering simpler preservation methods like cold static storage (CSS). This study provides a fundamental mechanistic comparison of myocardial injury pathways in human DBD and DCD hearts during cold storage, deliberately employing cold normal saline to isolate the exclusive impact of warm ischemia without the confounding protective effects of advanced preservation solutions. Our data reveal a stark divergence primarily driven by donor type (DBD vs. DCD). The DBD hearts demonstrate remarkable resilience to cold storage, whereas DCD hearts undergo a rapid, fWIT-dependent deterioration characterized by a failure of membrane repair and the activation of inflammatory pyroptotic cell death.

The core finding of this study is the identification of Annexin A1 depletion as the pivotal early event that distinguishes DCD from DBD hearts. We demonstrate that even prior to cold storage, DCD hearts exhibit significantly reduced levels of Annexin A1, a key protein essential for plasma membrane repair by promoting vesicle aggregation and fusion at injury sites.(12) This deficit was directly correlated with the duration of fWIT, underscoring warm ischemia as the primary insult. In contrast, DBD hearts, spared from fWIT, maintained stable and robust Annexin A1 expression throughout the 6-hour cold storage period. This finding is critical as plasma membrane integrity is the final barrier between reversible and irreversible cell injury.(8, 13) The rapid depletion of Annexin A1 in DCD hearts signifies a catastrophic failure of this first line of defense, rendering the cells profoundly vulnerable to subsequent insults, a vulnerability that is unmasked and exacerbated by the additional stress of cold storage.

The consequence of this membrane repair failure is the activation of a robust and irreversible inflammatory cell death pathway: pyroptosis. Our results clearly show that the NLRP3/caspase-1/GSDMD-NT axis, the canonical pathway for pyroptosis,(14) is significantly activated in DCD hearts at baseline and escalates dramatically during cold storage. The executioner protein, cleaved GSDMD-NT, forms pores in the plasma membrane, leading to cell rupture and the release of pro-inflammatory cytokines. Our previous study showed that myocardial inflammation is a preexisting condition in DCD hearts, where we did not observe any sign of inflammation in DBD hearts under cold storage.(10) The present study showed the catastrophic rise in TUNEL-positive cells in DCD hearts, exceeding 50%, compared to their near-absence in DBD hearts. This progression from ‘Annexin A1 depletion’ to ‘NLRP3 inflammasome activation’ to ‘caspase-1 cleavage’ to ‘GSDMD-NT pore formation’ to pyroptotic cell death provides a coherent and mechanistic explanation for the accelerated and irreversible damage observed in DCD hearts during cold storage. Importantly, the stability of these markers in DBD hearts confirms that cold storage itself is well-tolerated in the absence of a prior warm ischemic insult. The potential mechanism of ischemia injury induced myocardial membrane disruption and cardiac injury in human DCD hearts compared to DBD hearts is depicted in **Figure 7**.

**Figure 7.**
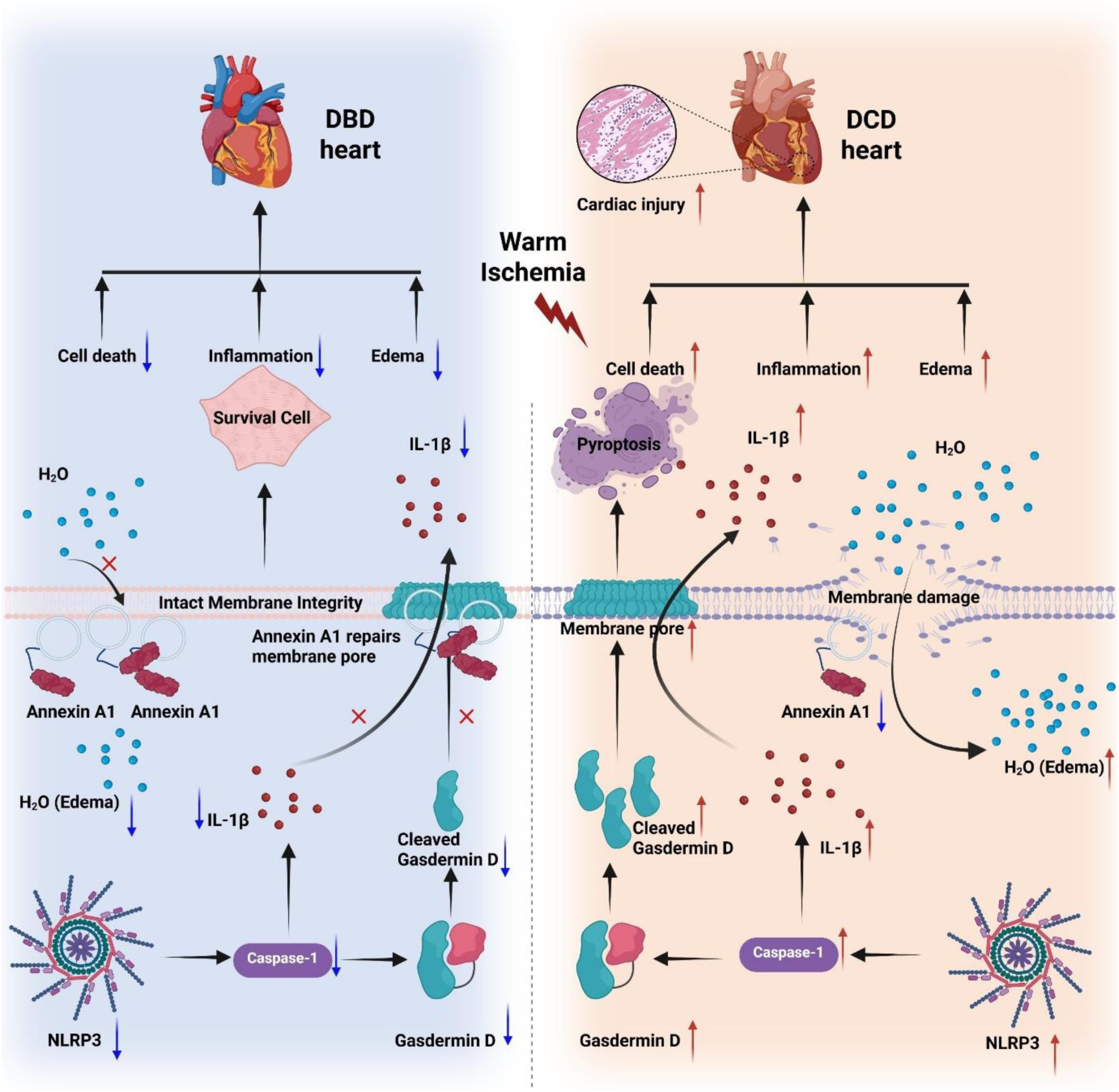
Mechanism of impaired membrane repair and myocardial damage induced by warm ischemia during cold storage of DCD hearts compared to DBD hearts. Created in BioRender. LI, S. (2025) https://BioRender.com/nfxtrko

A notable secondary finding was the differential role of Dysferlin, another membrane repair protein. Unlike Annexin A1, Dysferlin expression did not differ between DBD and DCD groups at baseline and our heatmap showed no correlation with fWIT or injury markers, suggesting it plays a minimal role in the initial response to fWIT. This aligns with mixed reports in the literature. One study showed that overexpression of Dysferlin has been shown to reduce cell death and alleviate hepatic ischemia injury.(15) However, another study by Han et al. using a coronary artery ligation model in Dysferlin knock-out mice found no significant differences in mortality, infarct size, and ejection fraction compared to wild-type mice, suggesting that Dysferlin does not confer protection against ischemic cardiomyopathy.(16) In another study by Tzeng et.al reported that Dysferlin mitigates myocardial ischemia/reperfusion injury.(17) In their study, the mice were subject to 30 minutes ischemia injury followed by 60 minutes reperfusion injury; there were no differences between wild-type mice and Dysferlin knock-out mice in cardiac function parameters like left ventricular developed pressure in the initial ischemia phase. Significant differences in cardiac function were observed during the reperfusion phase.(17) Consistent with these findings, we did not observe significant differences in cardiac Dysferlin expression during the initial warm ischemia phase. However, in DCD hearts, Dysferlin expression was significantly downregulated after four hours of cold static storage. This may suggest that Dysferlin is less sensitive to early ischemic injury than Annexin A1 and may participate in membrane repair after more extensive damage has occurred, but its value as an early biomarker or therapeutic target appears limited compared to Annexin A1.

The deliberate use of normal saline, rather than a specialized preservation solution, is a key strength of this study’s design. While solutions like Celsior, UW, or HTK offer cardioprotection and are the clinical standard, they introduce variables that can mask the fundamental injury mechanisms we sought to isolate. By using normal saline, we have effectively created a "worst-case scenario" model that reveals the true, unmitigated vulnerability of donor hearts to cold storage. This approach provides a clear baseline of the underlying pathophysiology, specifically, the Annexin A1-pyroptosis axis, that any future preservation strategy must overcome. Consequently, our findings are not a critique of cold storage per se, but rather a mechanistic roadmap for optimizing it. We propose that Annexin A1 levels and pyroptosis markers serve as crucial biomarkers for assessing donor heart quality during the fWIT phase and for evaluating the efficacy of future preservation interventions.

The translational implications of this work are substantial. For clinical practice in resource-limited settings or for short-distance transport, our findings suggest that simply using cold storage with normal saline is inadequate for DCD hearts. However, this study lays the groundwork for targeted therapeutic strategies. The strong negative correlation between Annexin A1 and injury suggests that its pharmacological supplementation, e.g., using Annexin A1 mimetic peptides, which have shown efficacy in ischemia-reperfusion models,(18–20) could be a promising strategy to bolster membrane repair in DCD hearts. Furthermore, inhibiting the pyroptosis pathway with NLRP3 or caspase-1 inhibitors(12) during procurement and preservation could synergistically protect these vulnerable organs. Future studies must now test these interventions, using the biomarkers established here, in conjunction with advanced preservation solutions or machine perfusion systems to determine if they can bridge the resilience gap between DBD and DCD hearts.

### Limitations

While this study provides novel mechanistic insights into the differential response of DBD and DCD hearts to cold storage, several limitations must be acknowledged. ***First***, the sample size, though substantial for a study of this nature involving human hearts, remains relatively small (n=24), particularly for the DBD control group (n=6). This was inherent to the challenge of procuring human hearts for research, a constraint that has become more pronounced following the FDA approval of DCD heart transplantation, as the majority of optimal DCD donors are now allocated to clinical use. This limited the power for extensive subgroup analyses across the spectrum of warm ischemic times. ***Second***, the demographic composition of our donor cohort may influence the generalizability of our findings. The median donor age was 45 years, which is older than the typical ideal cardiac donor, and the DCD group was predominantly male. As both advanced age and female sex have been associated in some studies with altered tolerance to ischemic injury,(21, 22), demographic composition may have influenced our results. However, this profile accurately reflects the "marginal donor" population that is increasingly considered for transplantation, thus enhancing the clinical relevance of our findings to real-world scenarios. ***Finally***, the use of cold normal saline instead of a specialized preservation solution was a deliberate choice to isolate the fundamental pathophysiological responses to warm and cold ischemia without the confounding protection of advanced additives. We acknowledge that this does not represent the current clinical standard of care for heart preservation. However, this approach was essential to establish a foundational understanding of the unmitigated injury mechanisms, specifically, the Annexin A1-pyroptosis axis, that any future preservation strategy must address. Consequently, our findings should be interpreted as revealing the underlying vulnerability of DCD hearts rather than as a direct critique of cold storage itself. We propose that the biomarkers identified herein serve as a crucial benchmark for evaluating the efficacy of optimized preservation solutions or machine perfusion systems in future studies. Despite these limitations, we believe the clear and consistent mechanistic pathways identified across multiple analytical modalities provide compelling evidence for the central role of Annexin A1 and pyroptosis in determining donor heart viability.

### Conclusions

In conclusion, this study identifies a fundamental mechanistic dichotomy between DBD and DCD hearts during cold storage. DBD hearts, protected from warm ischemia, are inherently resilient and tolerate cold storage in normal saline well. In stark contrast, DCD hearts suffer a double-hit injury, warm ischemia causes rapid Annexin A1 depletion, crippling membrane repair capacity, and priming the heart for a cascade of inflammatory pyroptotic cell death that is severely exacerbated by subsequent cold storage. The severity of this injury is directly proportional to the warm ischemic time. By utilizing normal saline, we have stripped preservation down to its most basic form, unequivocally revealing this vulnerable pathway and establishing Annexin A1 and pyroptosis markers as critical biomarkers of injury. This work provides a foundational mechanistic understanding that is essential for developing targeted strategies to repair the damaged DCD heart, ultimately aiming to increase the availability of viable donor hearts for transplantation.

## DATA AVAILABILITY

Data will be made available upon reasonable request.

## ACKNOWLEDGMENTS

We would like to express our sincere gratitude to Life Gift, the local organ procurement organization in Houston, Texas, for supplying the donor hearts used in this research. We also extend our heartfelt thanks to the donor’s next of kin for their generosity in allowing their loved one’s organ to be utilized for this important and innovative purpose. Lastly, we appreciate the Valour Foundation for their generous support through research funding.

## AUTHOR CONTRIBUTIONS

S.L., and N.K.M. conceived and designed research; S.L., R.B., A.E.E., K.V.N., S.B.P., and N.K.M. performed experiments; S.L., K.V.N., A.M.H., and N.K.M. analyzed data; S.L., A.M.H., K.K.L., and N.K.M. interpreted results of experiments; S.L., R.B., K.V.N., A.M.H., and N.K.M. prepared figures; S.L., and N.K.M. drafted manuscript; S.L., R.B., A.E.E., K.V.N., A.M.H., S.B.P., A.M.H., C.H.-M., T.K.R., K.K.L., and N.K.M. edited, revised, and approved final version of manuscript.

## Abbreviations

DBD: donation after brain death
DCD: Donation after circulatory death
fWIT: Functional warm ischemic time
CSS: cold static storage
OCS: Organ Care System
FDA: food and drug administration
GSDMD: gasdermin D
GSDMD-NT: N-terminal fragment of gasdermin D
NLRP3: NACHT, LPR, and PYD domains-Containing protein 3
TUNEL: Tdt-mediated dUTP nick end labeling

